# PBPK-led guidance for cystic fibrosis patients taking elexacaftor-tezacaftor-ivacaftor with nirmatrelvir-ritonavir for the treatment of COVID-19

**DOI:** 10.1101/2022.01.20.22269253

**Authors:** E. Hong, L.M. Almond, P.S. Chung, A.P. Rao, P.M. Beringer

## Abstract

**Background:** Cystic fibrosis transmembrane conductance regulator (CFTR) modulating therapies including elexacaftor, tezacaftor, and ivacaftor (ETI) are primarily eliminated through cytochrome P450 (CYP) 3A-mediated metabolism. This creates a therapeutic challenge to the treatment of COVID-19 with nirmatrelvir-ritonavir in people with cystic fibrosis (pwCF) due to the potential for significant drug-drug interactions (DDI). However, pwCF are more at risk of serious illness following COVID-19 infection and hence it is important to manage the DDI risk and provide treatment options.

**Methods:** CYP3A-mediated DDI of ETI was evaluated using a physiologically based pharmacokinetic (PBPK) modeling approach. Modeling was performed incorporating physiological information and drug dependent parameters of ETI to predict the effect of ritonavir (the CYP3A4 inhibiting component of the combination) on pharmacokinetics of ETI. The ETI models were verified using independent clinical pharmacokinetic and DDI data of ETI with a range of CYP3A modulators.

**Results:** When ritonavir was administered on day 1 through 5, the predicted AUC ratio of ivacaftor (the most sensitive CYP3A substrate) on day 6 was 9.31, indicating that its metabolism was strongly inhibited. Based on the predicted DDI, the dose of ETI should be reduced when co-administered with nirmatrelvir-ritonavir to elexacaftor 200mg-tezacaftor 100mg-ivacaftor 150mg on days 1 and 5, with resumption of full dose ETI on day 9, considering the residual inhibitory effect of ritonavir as a mechanism-based inhibitor.

**Conclusions:** Coadministration of nirmatrelvir-ritonavir requires a significant reduction in the ETI dosing frequency with delayed resumption of full dose due to the mechanism-based inhibition with ritonavir.

## 1. Introduction

The introduction of the Cystic Fibrosis Transmembrane Conductance Regular (CFTR) modulator, a triple combination of elexacaftor, tezacaftor, and ivacaftor (ETI, TRIKAFTA®) has resulted in significant improvements in lung function and nutritional status in people with cystic fibrosis (pwCF)(1). While ETI is indicated in up to 90% of the CF population(1), all 3 components are eliminated mainly through cytochrome P450 (CYP) 3A-mediated hepatic metabolism(2), and therefore present a therapeutic challenge in pwCF due to the potential for significant drug-drug interactions (DDI). The use of strong CYP3A inducers will increase the metabolism of ETI resulting in reduced exposure and a potential lack of efficacy, while concomitant therapy with agents that inhibit CYP3A will increase ETI levels placing the patient at increased risk of dose-related adverse effects. Therefore, the safe and effective use of CFTR modulators requires appropriate DDI management with concomitant CF medications.

One notable therapeutic challenge is in the treatment of COVID-19 (SARS-CoV-2). In pwCF, viral respiratory tract infections can lead to acute pulmonary exacerbations with a negative impact on lung function(3). COVID-19 infection triggers a cytokine storm which can lead to the life-threatening respiratory distress syndrome, potentially putting CF population infected with COVID-19 be at high risk of serious illness(4). The U.S. Food and Drug Administration (FDA) has recently issued an emergency use authorization (EUA) for the use of the nirmatrelvir-ritonavir for the treatment of mild-to-moderate COVID-19. Nirmatrelvir-ritonavir treatment significantly reduces hospital admissions and deaths among people with COVID-19 who are at high risk of severe illness(5). Nirmatrelvir is co-administered with ritonavir, a CYP3A inhibitor, to boost nirmatrelvir concentrations to achieve therapeutic levels(5). However, due to the potent inhibition effect of ritonavir, it may increase plasma concentrations of drugs that are primarily metabolized by CYP3A. Therefore, co-administration of nirmatrelvir-ritonavir is contraindicated with drugs highly dependent on CYP3A for clearance and for which elevated concentrations are associated with serious and/or life-threatening reactions. Since all 3 components of ETI are eliminated mainly through CYP3A, nirmatrelvir-ritonavir is expected to exhibit a significant drug interaction with ETI. Thus, the use of nirmatrelvir-ritonavir in pwCF would require an adjusted dosing regimen of ETI to prevent increased plasma concentrations and potential adverse drug reactions. However, there is currently no clinical data available regarding the interactions of ETI with nirmatrelvir-ritonavir and no specific dosing guidelines have been established. Therefore, there is an urgent need for the proper guidance regarding the use of nirmatrelvir-ritonavir for pwCF to prevent progression of COVID-19 to severe disease.

This study aimed to investigate the magnitude of the drug interactions of ritonavir-ETI, to simulate possible treatment scenarios and provide dosing recommendations to overcome the interaction. The CYP3A inhibition-mediated drug interaction of ETI was evaluated using a physiologically based pharmacokinetic (PBPK) simulation-based approach. PBPK simulation is a tool to predict the pharmacokinetic behavior of drugs in humans by integrating the information from multiple *in vitro* and clinical studies, exploring the effects of drug (e.g., physicochemical properties) and system (e.g., physiological) information on drug exposure. The predictive performance of PBPK simulations for CYP enzyme-based DDIs has been well established(6, 7), and this strategy is increasingly included during regulatory review by the FDA as an alternative for exploring DDI potential to provide dosing recommendations in product labeling(8). The present study contributes to improved treatment for COVID-19 in pwCF, by providing tools to evaluate and potentially overcome clinically important drug interactions involving highly active CFTR modulator therapy.

## 2. Methods

The workflow adopted for PBPK model development, verification, and application are illustrated in **Figure 1**. The models were implemented within the Simcyp Simulator (version 19; Certara, Sheffield, UK).

**Figure 1.**
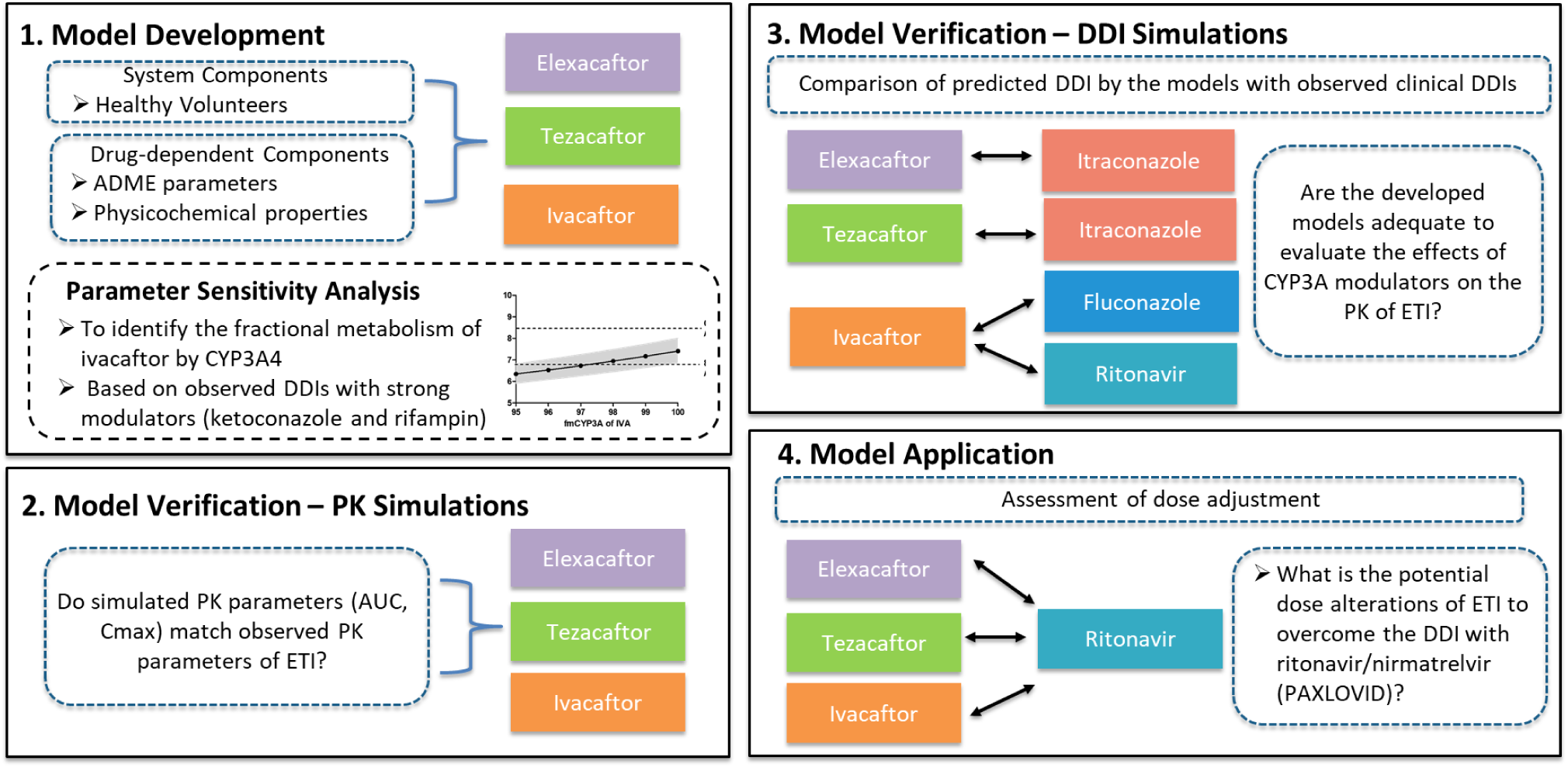
PBPK modeling framework detailing the processes of model development and verification that were performed in this study. Successful model verification must precede the application of the PBPK model of ETI for predictions of drug interactions with nirmatrelvir-ritonavir.

### 2.1. Model Development

#### PBPK Models of ETI

In the default healthy population library file (Sim-Healthy volunteers) provided in Simcyp^®^, the distribution of ages and proportion of female were corrected to reflect the demographics of CF population based on patient registry 2020 annual report published by cystic fibrosis foundation(9). Specifically, the frequency of population aged 18-21 years, was adjusted from 4.5% in healthy population to 13.1% in CF. Also, the proportion of females was adjusted from 0.32 in healthy population to 0.48 in CF. The mean body mass index (BMI) of healthy population (23.5 kg/m2) was similar to that observed in CF (21.2 kg/m2 in 2005 to 23.1 kg/m2 in 2020) and no further adjustment was needed, reflecting how BMI in pwCF has been increased over the years with continued improvements in CF care (9, 10). All other system parameters were kept as the default healthy, and this assumption is in line with the PK parameters of ETI not differing between healthy adults and patients with CF(11-13). This population library file was used for all simulations. For trial design, we used a total size of 100 population (10 trials and 10 subjects in each trial). The PBPK model input parameters for ETI are summarized in **Table S1-S3** and described in detail below. The ivacaftor model consists of the advanced dissolution, absorption, and metabolism (ADAM) model and minimal PBPK model. The ADAM model divides the gastrointestinal tract into nine segments, where the drug absorption from each segment is predicted by a function of drug dissolution, luminal degradation, metabolism, transport, etc(14). The minimal PBPK model consists of no more than five compartments, that include the gastrointestinal tract, blood, liver, and up to two additional compartments, reducing the complexity of the model while allowing for mechanistic simulations in the compartment of interest(15). The ivacaftor model was constructed based on available physicochemical properties and clinical data from published PK studies(11, 16-19). Ivacaftor is a diprotic acid with an acid dissociation constant (pKa) of 9.4 and 11.6. Ivacaftor has a logP of 5.68, and predominantly binds to albumin with fraction of unbound drug in plasma of 0.001. The blood/plasma partition ratio was 0.55. The absorption parameter *P*_eff,man_ was predicted by the Simcyp calculator based on its physicochemical properties. The distribution parameters including *V*_sac_ and *V*_ss_ were obtained from published data. The *in vitro* studies and clinical DDI data suggest that ivacaftor is predominantly eliminated through CYP3A4 mediated hepatic metabolism(13). Therefore, the excretion was set to enzyme kinetics to quantify its metabolism by CYP3A. The intrinsic clearance by CYP3A4 was back calculated from the oral clearance observed in healthy subjects (19.0 L/h)(11). The fraction of ivacaftor being metabolized by CYP3A4 (fmCYP3A4) was set to 98% in order to capture the observed drug interactions of ivacaftor with the strong CYP3A4 modulators, ketoconazole or rifampin. The *F*_g_ was also optimized to 0.50 using observed DDI data. The additional human liver microsomes (HLM) clearance representing non-CYP3A pathways was also estimated using the retrograde calculator and entered into the model to capture the observed pharmacokinetics profiles.

The tezacaftor and elexacaftor PBPK models were constructed based on the data from the PBPK review section within the NDA documents and the publication of Tsai et al(12, 13, 17). Briefly, the elexacaftor and tezacaftor models consist of the first-order absorption and a minimal PBPK model. In the NDA document, it was described that the absorption and distribution parameters were obtained from the observed PK profiles following clinical phase 1-3 studies, and the fmCYP3A4 of elexacaftor and tezacaftor were set to 67% and 73.2% based on human ADME studies. Using the retrograde model, the rCYP3A4 CL_int_ values of elexacaftor and tezacaftor attributed to CYP3A4 were calculated to be 0.233 and 0.175 μL/min per picomole of isoform. The M1-tezacaftor, active metabolite of tezacaftor, was also incorporated into the tezacaftor PBPK model based on data in the NDA document(12). The other active metabolites, M23-elexacaftor and M1-ivacaftor, were not incorporated into the PBPK analyses due to insufficient information to build the model.

#### PBPK Models of CYP3A Modulators

Rifampin, ketoconazole, fluconazole, itraconazole, and its primary metabolite, hydroxy itraconazole, are prototypical CYP3A modulators that have been implicated in clinical drug interaction studies with elexacaftor, tezacaftor, or ivacaftor. Ritonavir is also a CYP3A modulator that we aimed to predict interactions with ETI. For simulation of DDIs, the validated compound files of these CYP3A modulators provided in Simcyp**®** (version 19) were used.

### 2.2. Model Verification: PK Simulations

The PK profiles of ETI following a single oral dose administration and multiple administrations of clinically relevant doses (elexacaftor 200mg qd, tezacaftor 100mg qd, and ivacaftor 150mg q12h) were first simulated to verify the performance of the PBPK models. ETI was orally administered under fed conditions to mimic the clinical setting, where the fat-containing food is required for optimal absorption of ETI. The simulated data were qualified using the observed PK data in a CF population aged older than 17 years old. The prediction accuracy for the area under the curve (AUC) and maximum plasma concentration (C_max_) values were calculated as a ratio of mean observed values over mean predicted values. Successful model performance was defined by mean ratios of AUC and *C*_max_ within a two-fold range as previously described(20, 21).

### 2.3. Model Verification: DDI Simulations

Upon accurate recapitulation of ETI’s PK, the models were further assessed against the clinical DDI data to verify fmCYP3A4 and establish if the models were adequate for the assessment of victim DDI liability. For verification simulations, the dose and schedule of drugs were matched to the design of the corresponding clinical DDI study in healthy subjects. For elexacaftor, itraconazole solution (200 mg) was administered daily from Day 1 to Day 10 and a single 20 mg dose of elexacaftor was administered orally on Day 5(13). For tezacaftor, the itraconazole solution (200 mg) was administered BID on Day 1 and QD from Day 2 to Day 14 and tezacaftor 25 mg was administered daily from Day 1 to Day 14(12). For ivacaftor, four clinical DDI studies were conducted with ritonavir, ketoconazole, fluconazole, or rifampin(11, 22). For ritonavir, 50mg was administered daily from Day 1 to Day 18 and a single 150 mg dose of ivacaftor was administered on Day 15. The ketoconazole 400 mg was administered daily from Day 1 to Day 10 and a single 150 mg dose of ivacaftor was administered on Day 4. For fluconazole, 400mg was administered on Day 1 and 200mg was administered daily from Day 2 to Day 8, and ivacaftor 150 mg was administered BID from Day 1 to Day 8. For rifampin, 600mg was administered daily from Day 1 to Day 10 and a single 150 mg dose of ivacaftor was administered on Day 6. To quantify the DDIs, the geometric mean ratios of AUC or C_max_ with or without the presence of CYP3A4 modulators were calculated. The assessment of DDI prediction success was based on whether predictions fall within a two-fold range of the observed data.

### 2.4. Model Application: DDI Predictions of ETI with Ritonavir

Although ritonavir-nirmatrelvir (PAXLOVID) is a fixed dose combination of 2 drugs, nirmatrelvir has not been included in simulations as there is no clinical evidence that it modulates CYP3A4 activity, and we have focused on interactions with ritonavir which is a potent CYP3A4 inhibitor. The verified PBPK-DDI model was applied to (1) predict the effect of ritonavir on the PK of ETI and (2) determine a potential dose alteration of ETI to overcome the CYP3A inhibition mediated by ritonavir. We first simulated the steady-state PK of standard dose ETI alone and when co-administered with 100mg ritonavir twice daily for 5 days based on the instruction for dosage and administration in the FDA-approved fact sheet of nirmatrelvir-ritonavir (5). In addition, since ritonavir acts as a mechanism-based CYP3A4 inhibitor by covalently binding to the CYP3A4(23), simulations were run until CYP3A4 and the PK of ETI had returned to baseline (10 days after ritonavir discontinuation). We then simulated several adjusted dosing regimens of ETI when co-administered with ritonavir to find the regimen that could provide the closest PK profiles of standard dosing of ETI alone.

## 3. Results

### 3.1. Development and Verification of the Models of ETI

#### 1) Parameter Sensitivity Analysis of the Fractional Metabolism of Ivacaftor by CYP3A4 on the Predicted AUC Ratio with CYP3A Modulators

For the PBPK model of ivacaftor, in the absence of an *in vitro* estimate, clinical interaction data with strong modulators of CYP3A4 were used to assign fmCYP3A4. First, we predicted DDI with ketoconazole by varying the fmCYP3A4 value of ivacaftor from 95% to 100% (**Figure S1A**), since it has been reported that the fractional metabolism of ivacaftor assigned to CYP3A4 is greater than 95%(17). An fmCYP3A4 of 98 % predicted the AUC ratio (GMR 6.95) of ivacaftor within the bioequivalence limit (80-125%) of the observed AUC ratio (GMR 8.45). Repeating this analysis for rifampicin, indicated the magnitude of DDI was well captured with fmCYP3A4 between 98 and 100% (simulated GMR 0.11 vs. observed 0.11) (**Figure S2B**). Taken together, the fmCYP3A4 value of 98% was chosen as it describes observed DDIs between ivacaftor and ketoconazole or rifampin within the bioequivalence limit.

#### 2) PBPK Models of ETI Recapitulated Clinically Observed PK Profiles

Model predictive performance of ETI was assessed using observed pharmacokinetic data sets from clinical trials(12, 13). The observed and simulated plasma concentration-time profiles of ETI following a single oral dose administration of elexacaftor 200mg, tezacaftor 100mg, and ivacaftor 100mg in healthy subjects are shown in **Figure S2**. For elexacaftor and tezacaftor, the mean plasma concentrations were used in the graph while the median plasma concentrations were used for ivacaftor as median value was reported from ivacaftor single dose PK study. The pharmacokinetic profile of ETI after single oral dose administration was captured by the PBPK model.

Further verification of the model was performed by simulating the steady-state PK of ETI when administered with multiple oral doses of elexacaftor 200mg qd, tezacaftor 100mg qd, and ivacaftor 150mg q12h. The predicted steady-state AUC and *C*_max_ of ETI were in the range of 0.9 to 1.2 of the observed values demonstrating the excellent performance of the model. The observed and simulated PK parameters of ETI are summarized in **Table 1**.

**Table 1.**
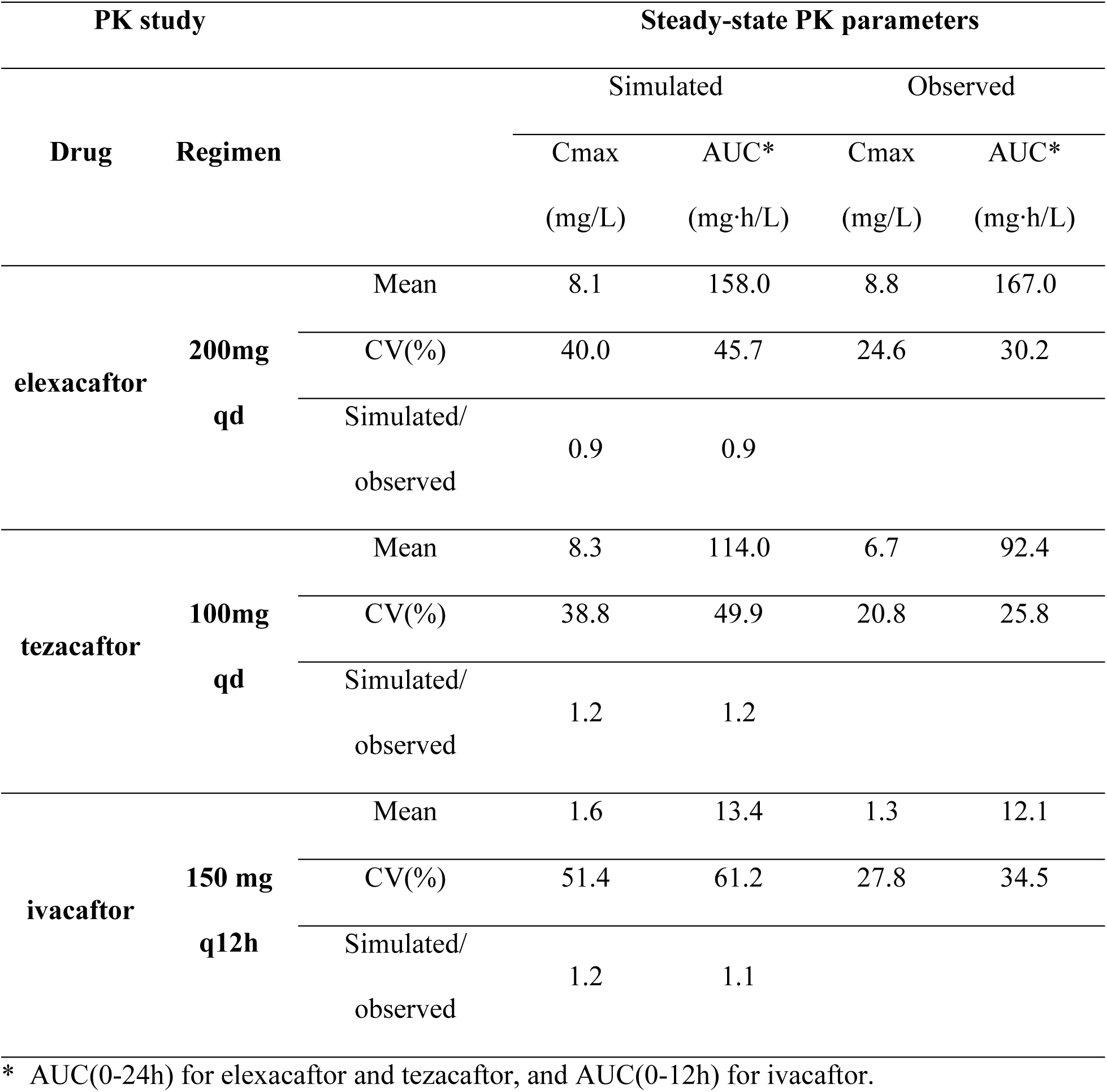
Comparison of PK parameters between simulated and observed data for model verification of ETI

#### 3) PBPK-DDI Models of ETI Recapitulated Clinically Observed Drug Interactions

Although preliminary PK simulations verified the predicted PK ETI, given that the PBPK models of ETI are intended to be applied for the characterization of DDIs involving CYP3A modulation, it is essential to verify the victim properties defined in the models by simulating independent clinical DDI studies with a range of perpetrator drugs. The robustness of the model was assessed by comparing the magnitude of simulated DDIs of ETI with that observed from the clinical trials. The PBPK-DDI models accurately recapitulated the observed DDI magnitude (**Table 2**).

**Table 2.** Summary of the simulated vs. observed Geometric Mean Ratio (GMR) of PK parameters in the presence and absence of CYP3A modulators

| DDI study | PK Parameters | Simulated<br>GMR (90% CI) | Observed<br>GMR (90% CI) | Ratio<br>(Simulated/<br>Observed) |
| --- | --- | --- | --- | --- |
| Ivacaftor | Cmax Ratio | 2.61 (2.47, 2.77) | 2.28 (1.84, 2.83) | 1.14 |
| +/- Ritonavir | AUC Ratio | 3.64 (3.37, 3.94) | 3.06 (2.36, 3.97) | 1.19 |
| Ivacaftor | Cmax Ratio | 2.04 (1.96, 2.12) | 2.65 (2.21, 3.18) | 0.77 |
| +/- Ketoconazole | AUC Ratio | 6.95 (6.44, 7.49) | 8.45 (7.14, 10.01) | 0.82 |
| Ivacaftor | Cmax Ratio | 2.72 (2.64, 2.81) | 2.47 (1.93, 3.17) | 1.10 |
| +/- Fluconazole | AUC Ratio | 3.33 (3.21, 3.46) | 2.95 (2.27, 3.82) | 1.13 |
| Ivacaftor | Cmax Ratio | 0.29 (0.27, 0.31) | 0.20 (0.17, 0.24) | 1.45 |
| +/- Rifampin | AUC Ratio | 0.11 (0.10, 0.13) | 0.11 (0.10, 0.14) | 1.00 |
| Tezacaftor | Cmax Ratio | 2.62 (2.53, 2.72) | 2.83 (2.62, 3.07) | 0.93 |
| +/- Itraconazole | AUC Ratio | 3.85 (3.65, 4.07) | 4.02 (3.71, 4.63) | 0.96 |
| Elexacaftor | Cmax Ratio | 1.08 (1.08, 1.09) | 1.05 (0.98, 1.13) | 1.03 |
| +/- Itraconazole | AUC Ratio | 2.00 (1.94, 2.06) | 2.83 (2.59, 3.10) | 0.71 |

### 3.2. DDI Simulation of ETI with Ritonavir

#### 1) Simulated DDI of ETI and Ritonavir Suggests Significant DDI with ETI

The verified PBPK-DDI models of ETI were used to simulate the standard dose of ETI when co-administered with nirmatrelvir/ritonavir to determine the magnitude of the DDI for its intended use for treatment of COVID-19. To mimic the clinical setting of nirmatrelvir/ritonavir administration, we simulated steady state PK of ETI, then ritonavir q12h for 5 days while continuing ETI standard dosing during and after ritonavir administrations. We calculated the *C*_max_ and AUC ratio of ETI in the presence and absence of ritonavir on day 6 of co-administration (**Table S4**). The magnitude of DDI achieves its maximum level on day 6, as ETI haven’t gotten to the steady-state after ritonavir administration, so drugs are still accumulating after the last dose of ritonavir (**Figure 2A, B, and C**). Further, the maximum CYP3A4 inhibition effect is maintained through day 6 (**Figure 2E**). The simulated geometric mean AUC ratio was highest for ivacaftor (9.31, 90% CI: 8.28, 10.47), followed by tezacaftor (3.11, 90% CI: 2.96, 3.27) and elexacaftor (2.31, 90% CI: 2.20, 2.42).

**Figure 2.**
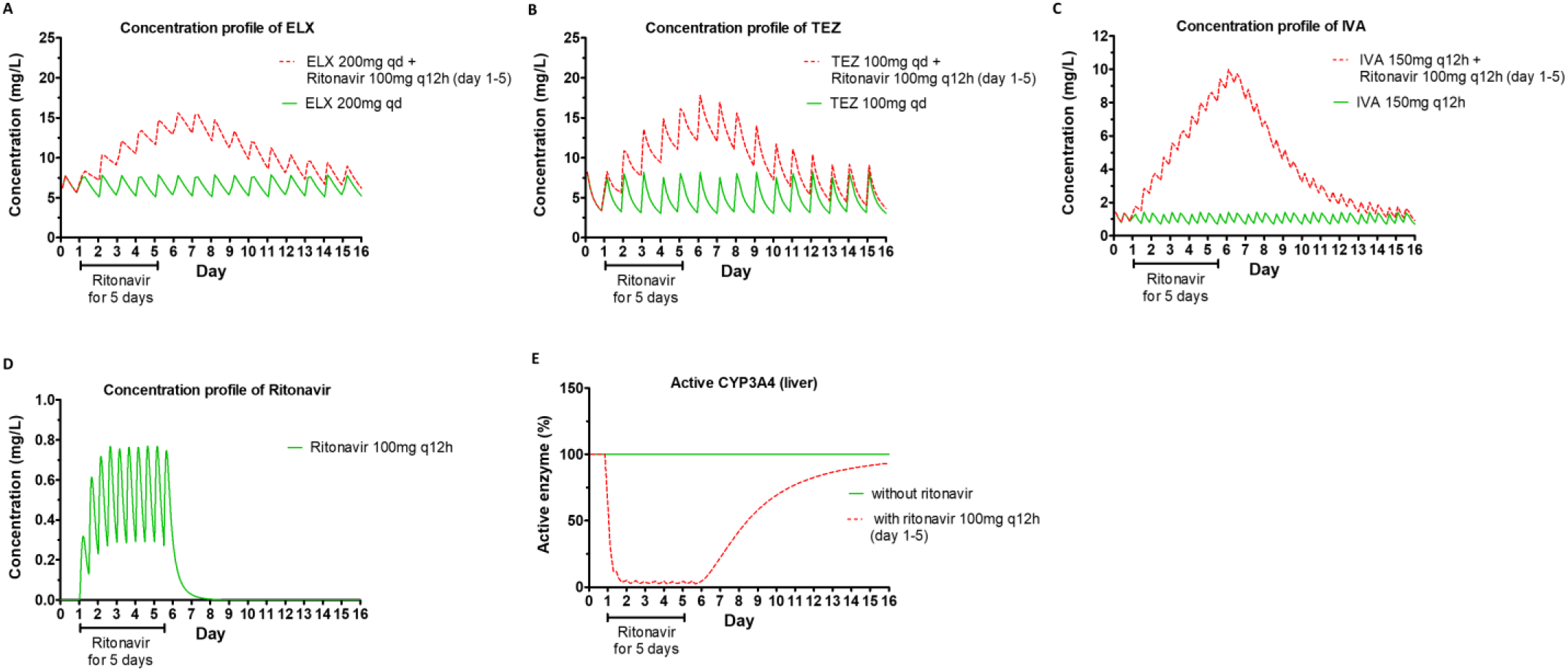
Plasma Concentration Profile of Elexacaftor (A), Tezacaftor (B), Ivacaftor (C), and Ritonavir (D), and the % of Active CYP3A4 Enzyme (E) Over Time. Green: without ritonavir, Red: with ritonavir administered day 1 through day 5.

Plasma concentrations of ETI in the presence and absence of ritonavir is shown in **Figure 2**. Although ritonavir itself is eliminated the day after discontinuation (**Figure 2D**), the CYP3A4 inhibition is time-dependent(24), so the inhibition is prolonged and the recovery to baseline time is reliant on the turnover of the CYP3A4 itself (**Figure 2E**). Thus, baseline steady state of all ETI drugs is predicted to be re-established on day 15. Crucially, this indicates that dose adjustment of ETI in case of co-administration with nirmatrelvir/ritonavir would be required to extend beyond the 5 days of co-administration.

#### 2) Altered Dose of ETI to Recapitulate the PK profile of Standard Dose ETI Alone

We next utilized the models to simulate ETI dose adjustments when these agents are co-administered with ritonavir and determine how long the adjusted dosage needed to be maintained, to overcome the enzyme inhibition effect mediated by ritonavir. Based on the simulated effects of ritonavir, elexacaftor 200mg, tezacaftor 100mg, ivacaftor 150mg in the morning (2 orange tablets) every 4 days (administered on day 1 and day 5 and resumed full dose on day 9) provided similar steady-state PK profile of the conventional regimen of ETI alone (**Figure 3**). The trough concentrations of ETI were all above the EC50 targets, which are 0.99 mg/L, 0.5 mg/L, and 0.048 mg/L for elexacaftor, tezacaftor, and ivacaftor, respectively(11-13). Since CYP3A4 inhibition dynamics mediated by ritonavir changes over time, we measured the mean *C*_max_ and AUC of reduced dosing of ETI regarding the first dose on day 1 and the second dose on day 5 and calculated the percentage of ETI standard regimen alone (**Table 3**). The AUC(0-96h) of reduced dosing regimen ranged from 83.0-142.5% of ETI alone. Resumption of the full dose of ETI on day 9 is based on simulations to optimize the concentration profiles of all components of ETI, where the level of elexacaftor and tezacaftor do not become lower than 80% of the standard regimen before resuming the full dose, while striving to maintain levels of ivacaftor below 125% of the standard regimen after resuming the full dose. At day 9, the CYP3A4 enzyme activities were recovered to 60% of the steady-state values.

**Figure 3.**
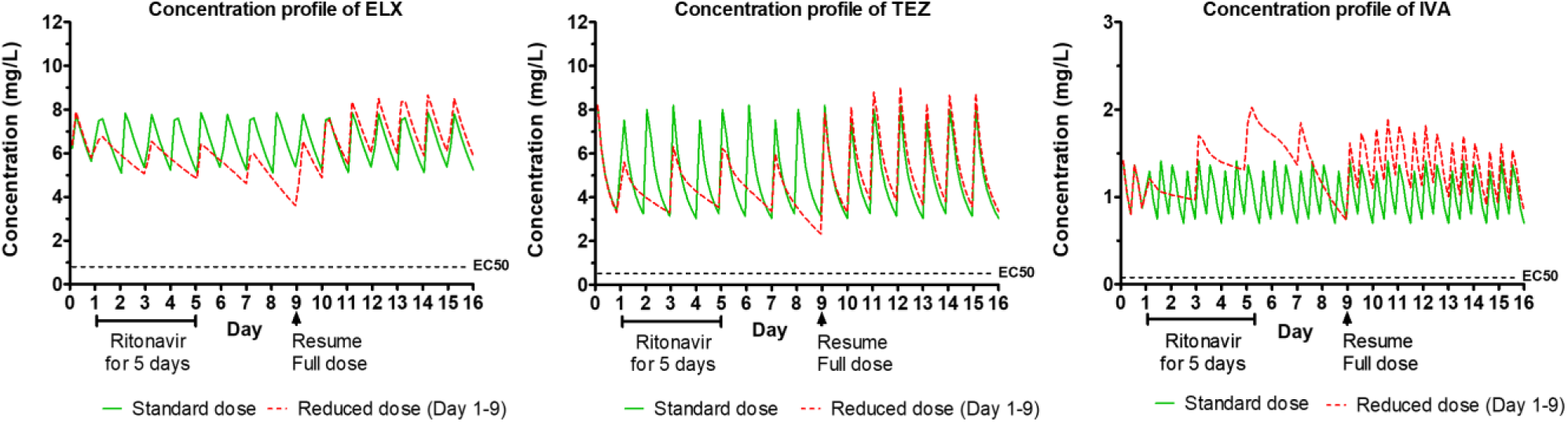
Plasma Concentration Profile of ETI. Green: standard dose without ritonavir, Red: reduced dose with ritonavir 150mg q12h administered day 1 through day 5. (EC50 for tezacaftor and ivacaftor: obtained from exposure-response analysis in clinical trials regarding the reduction of sweat chloride, EC50 for elexacaftor: obtained from in vitro study of chloride transport in phe508del/phe508del human bronchial epithelial cells as no in vivo data are available.)

**Table 3.**
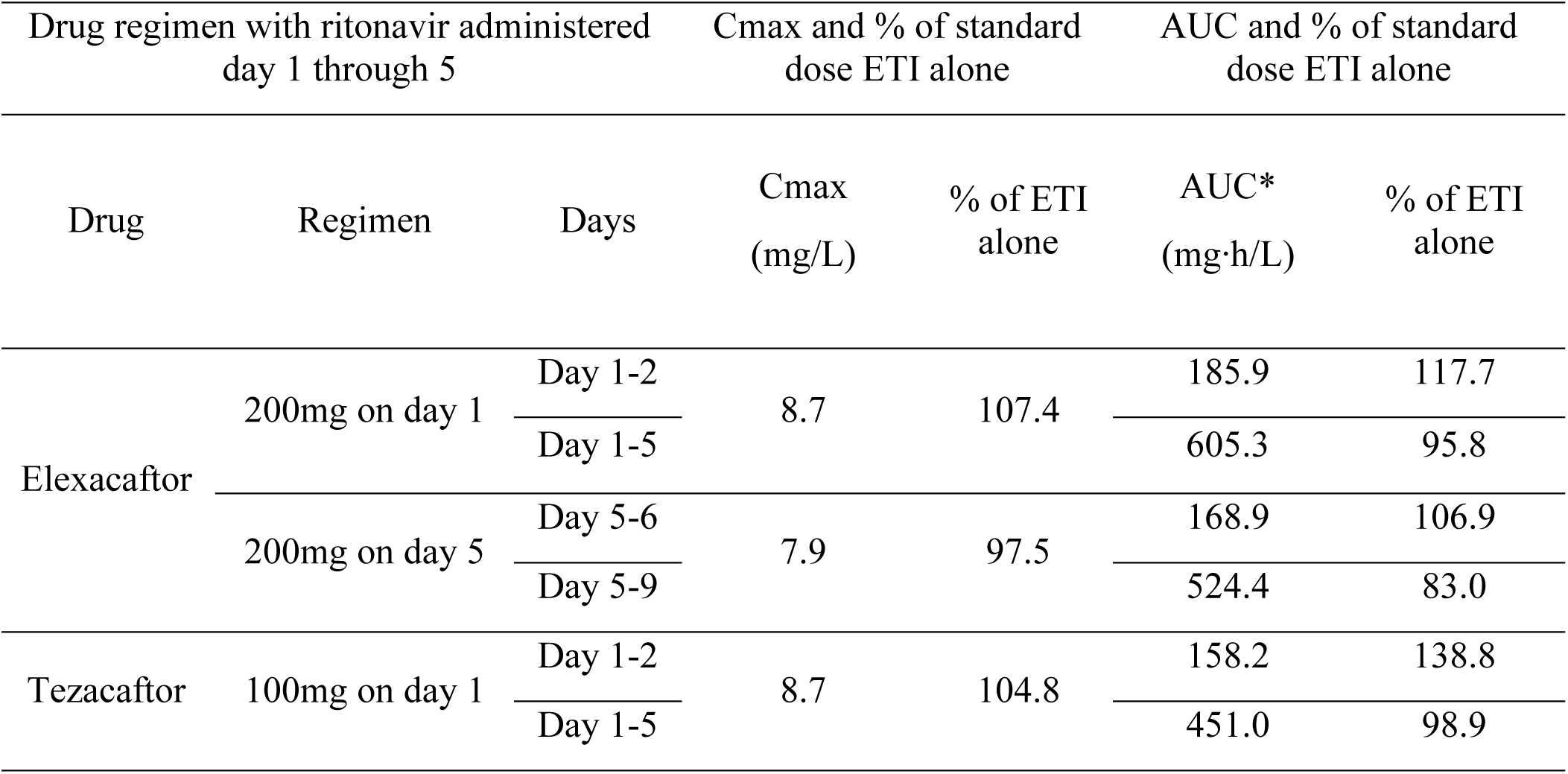
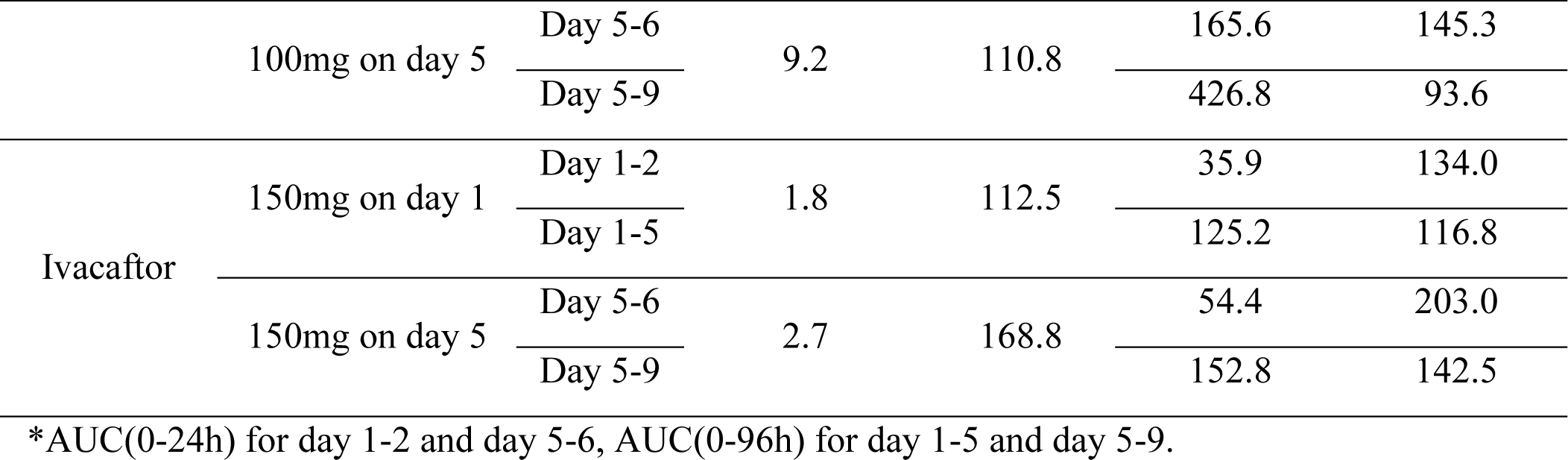
Predicted mean Cmax and AUC of reduced dose of ETI (two orange tablets q96h instead of two orange tablets in the morning and one blue tablet in the evening) with ritonavir 150mg q12h administered day 1 through day 5

In addition, we simulated an alternate dosing regimen, which is elexacaftor 100mg, tezacaftor 50mg, ivacaftor 75mg in the morning (1 orange tablet) administered every 2 days. This regimen provided concentration profiles closer to the standard regimen of ETI alone with less fluctuations of peak/trough concentrations (**Figure S3 and Table S5**). Especially for ivacaftor on day 5 which showed higher *C*_max_ (2.7 mg/L, 168.8% of standard regimen) in case of 150mg q96h, the *C*_max_ of ivacaftor was 2.1 mg/L (131.3 % of standard regimen) with the 75mg q48h. However, the dosing regimen of two orange tablets every 3-4 days is consistent with recommendations for other strong CYP3A inhibitors and the *C*_max_ and AUC values between the two regimens were not demonstrably different.

For the dosing recommendation of tezacaftor/ivacaftor (SYMDEKO), since it is provided as a fixed dose yellow tablet consisting of tezacaftor 100mg and ivacaftor 150mg, the same dosing recommendation above (tezacaftor-ivacaftor 100-150mg q96h) can be applied. Also, for ivacaftor 150mg tablet (KALYDECO), the same dosing recommendation (one tablet q96h) can be applied, but alternatively, the dosing interval of ivacaftor could be further increased to 5 days rather than 4 days, to recapitulate similar PK profile of standard regimen. When ivacaftor 150mg was administered on day 6 instead of day 5, the AUC(0-24h) was decreased to 48.31 mg·h/L (180.2% of standard regimen) from 54.4 mg·h/L (203.0% of standard regimen) (**Table S6**). Taken together, the suggested dosing schedule of CFTR modulators co-administered with nirmatrelvir/ritonavir is described in **Figure 4**.

**Figure 4.**
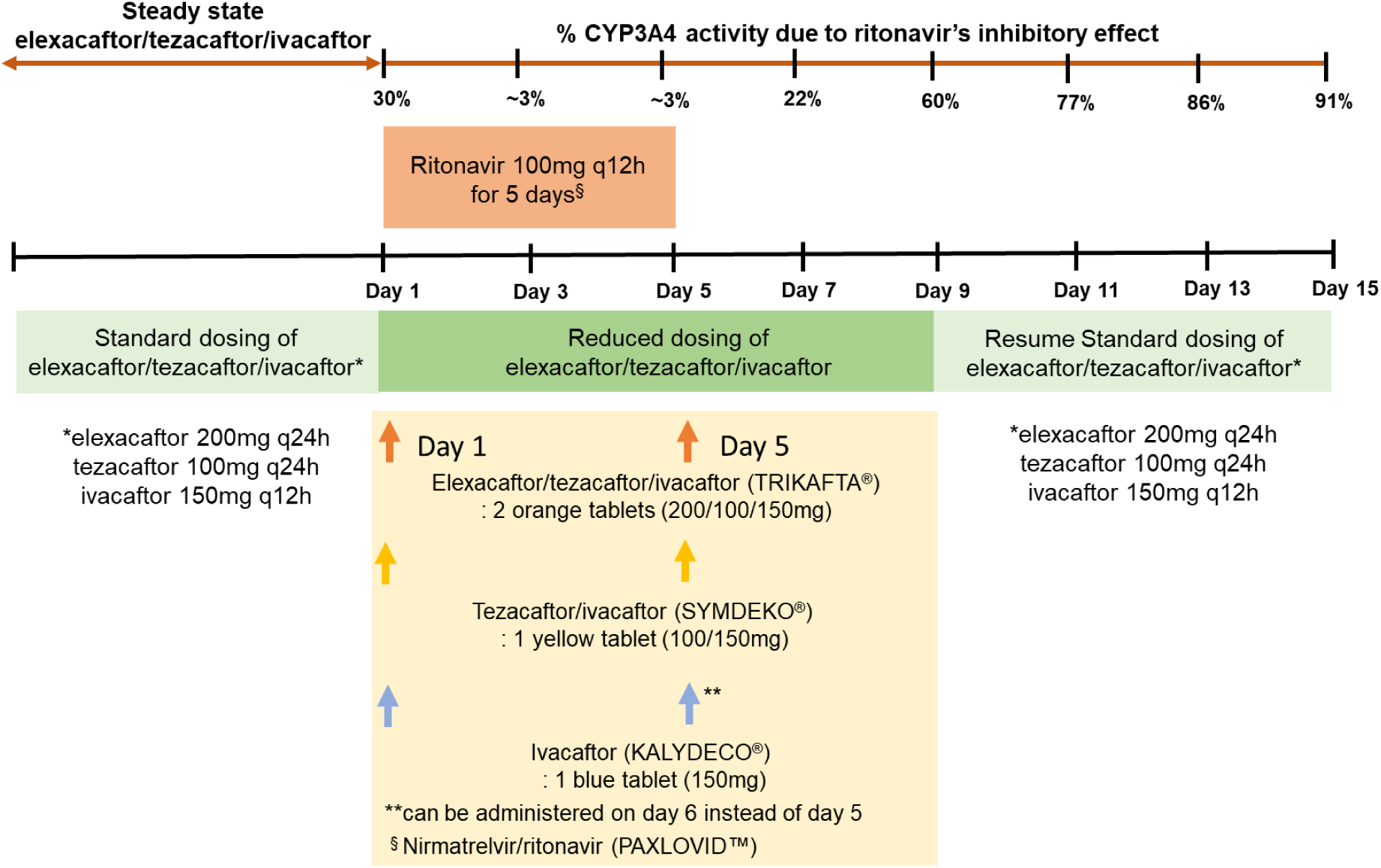
Suggested Dosing Schedule of CFTR modulators co-administered with nirmatrelvir-ritonavir

## 4. Discussion

All three components of ETI are eliminated predominantly through hepatic metabolism along with limited renal excretion. The clinical DDI study with strong CYP3A4 inhibitors (Ketoconazole and Itraconazole) or inducer (Rifampin) showed that ETI are the sensitive CYP3A4 substrates. Therefore, the safe and effective use of CFTR modulators is complicated by DDI management with concomitant CF medications, as CYP3A4 modulation by inducers or inhibitors can lead to altered systemic exposure, resulting in variability in drug response. CF patients often take multiple antibiotics including rifamycins, macrolides, and azole antifungals, which potentially inhibit or induce CYP3A4-mediated metabolism of ETI. Recently Tsai et. al. published a ETI PBPK model to evaluate exposures during the transition from mono or dual combination of CFTR modulators to ETI(17). We extended the models by refining the ivacaftor model and further validating the ETI PBPK-DDI model with published clinical DDI data. Since the ETI PBPK-DDI model we employed could robustly predict PK parameters and the observed drug interactions of ETI, it can provide an approach to the evaluation and management of other potential DDIs involving CFTR modulators.

In particular, we aimed to provide guidance for ETI dose adjustment with ritonavir, the CYP3A inhibitor and the component of nirmatrelvir/ritonavir for the treatment of COVID-19. From the ETI-ritonavir DDI simulations, we found that when ritonavir 100mg q12h was administered for 5 days, it led to the AUC ratio of ivacaftor as 9.31 (90% CI: 8.28, 10.47) which far exceeded the observed and simulated AUC ratio (3.06) when ivacaftor was administered with ritonavir 50 mg q24h(22). The increase in interaction with the therapeutic regimen shows that dose adjustments should not be estimated purely on the basis of the available clinical study. Further, through the simulations we found that the elevated concentrations of ETI were continued even after ritonavir is eliminated due to the irreversible CYP3A4 inhibition effect. The suggested reduced dosing regimen that is maintained until 4 days after ritonavir is discontinued provided a similar PK profile of the standard regimen of ETI alone. However, dose-dependent adverse reactions of ivacaftor should be more closely monitored due to its high peak concentrations on day 5. This study has important clinical implications, bridging the gap between the available clinical DDI study and that of the therapeutic regimen and providing proper dosing guidelines for treatment of COVID-19 with nirmatrelvir/ritonavir in pwCF receiving concomitant CFTR modulator therapy.

A limitation of this study is that in the absence of data, population system parameters, such as plasma protein levels in pwCF was not incorporated into the modelling. However, changes in demography reflecting CF population were incorporated. Furthermore, a prior study evaluating the hepatic clearance of drugs showing that CYP3A enzyme activity is unaffected in pwCF(25) and the current weight of evidence based on comparisons of ETI PK in healthy volunteers compared to patients suggests they are comparable(11-13). Previous studies indicate that differences in pharmacokinetics of drugs in CF is attributed to differences in body composition and plasma protein concentrations secondary to nutritional deficiencies(26). However, the BMI of pwCF has increased over the years with continued improvements in CF-care, including highly effective CF modulators and nutritional support(9), to the extent it is now similar to that of healthy volunteers(10).

A potential limitation of this work is that we did not include models of all active metabolites of ETI. M1-tezacaftor is an important metabolite due to its similar potency with parent drug as well as its high metabolite to parent AUC ratio (157.8%)(12). Since M1-tezacaftor is also metabolized by CYP3A4(12), its formation and elimination may be altered with ritonavir co-administration. Using the PBPK model of M1-tezacaftor, we were able to determine that the reduced dose of ETI provided mean AUC(0-96h) for M1-tezacaftor during the co-administration period with ritonavir that was 80.9% of the standard regimen of ETI alone. We did not include models of active metabolites of ivacaftor (M1-ivacaftor) and elexacaftor (M23-elexacaftor), since there was insufficient information to build the models incorporating them. While it is currently unknown whether ritonavir would alter the levels of M23-elexacaftor, the exposure of this metabolite is significantly reduced when compared with the parent compound and is not considered to contribute significantly to the overall efficacy(13). For M1-ivacaftor, there is evidence that its plasma concentration of is decreased by 35% in the presence of rifampin, indicating that M1-ivacaftor may also metabolized by CYP3A4(27). This perhaps suggests that the reduced dose of ivacaftor may not significantly reduce M1-ivacaftor exposure when co-administered with ritonavir, since ritonavir will inhibit the metabolism of both ivacaftor and M1-ivacaftor.

In conclusion, using a PBPK modeling approach, we demonstrated that nirmatrelvir/ritonavir can be administered concomitantly with ETI in pwCF with proper dose adjustment. The outcome of this study ensures the use of nirmatrelvir/ritonavir for the treatment of COVID-19 in patients with CF while continuing to receive highly active CFTR modulators. In addition, this work provides tools to evaluate and potentially overcome clinically important DDIs involving highly active CFTR modulator therapy.

## Data Availability

All data produced in the present work are contained in the manuscript.

