## Supplemental Tables and Figures for "PBPK-led guidance for cystic fibrosis patients taking elexacaftor-tezacaftor-ivacaftor with nirmatrelvir-ritonavir for the treatment of COVID-19"

**Table S1.** Parameters Used to Develop the Ivacaftor Model in Simcyp**®** version 19 (Certara)

| **Parameter** | **Value** | **Source** |
| --- | --- | --- |
| Physiochemical properties | |  |
| Molecular weight (g/mol) | 392.49 | CAS ID: 873054-44-5 |
| Log P_o:w_ | 5.68 | [1] |
| Compound type | Diprotic acid | [1] |
| p*K*_a_ 1 | 9.4 | [1] |
| p*K*_a_ 2 | 11.6 | [1] |
| B/P | 0.55 | [2] |
| fu_p_ | 0.001 | [2, 3] |
| Absorption | |  |
| Absorption model | ADAM |  |
| Caco-2 permeability (10^−06^ cm/sec) | 119 | [2] |
| fu_gut_ | 0.279 | optimized |
| *k*_a_ (h^−1^) | 5.188 | Predicted in Simcyp |
| *F*_g_ | 0.5 | optimized |
| *P*_eff,man_ (× 10^−4^ cm/s) | 11.9 | Predicted in Simcyp |
| Distribution | |  |
| Distribution model | Minimal PBPK model |  |
| *Q* (l/h) | 3.752 | [2] |
| *V*_sac_ (l/kg) | 6.70×10^−01^ | [2] |
| *V*_ss_ (l/kg) | 1.89 | [2] |
| Elimination | |  |
| CL_po_ (l/h) | 19.0 | [4, 5] |
| rCYP3A4 CL_int_ (µL/min/pmol of isoform) | 19.9 | Predicted in Simcyp using the Retrograde Calculator |
| Additional HLM clearance (µL/min/mg protein) | 78.8 | Predicted in Simcyp |

ADAM, advanced dissolution, absorption, and metabolism; B/P, blood-to-plasma ratio; CL_int_, intrinsic clearance; CL_po_, in vivo oral clearance; *F*_g_, fraction escaping gut-wall elimination; fu_gut_, fraction unbound in the enterocyte; fu_p_, fraction unbound in plasma; HLM, human liver microsome; *k*_a_, absorption rate constant; Log P_o:w_, logarithmic partition coefficient octonal:water; *P*_eff,man_, effective permeability in man; p*K*_a_, logarithm of acid dissociation constant; *Q*, inter-compartment clearance; *V*_sac_, single adjusted compartment volume; *V*_ss_, volume of distribution at steady state.

**Table S2.** Parameters Used to Develop the Tezacaftor Model in Simcyp**®** version 19 (Certara)

| **Parameter** | **Tezacaftor** | **M1-tezacaftor** | **Source** |
| --- | --- | --- | --- |
| Physiochemical properties | |  |  |
| Molecular weight (g/mol) | 520.5 | 518.48 | [2, 6] |
| Log P_o:w_ | 3.6 | 4.397 | [2, 6] |
| Compound type | Neutral | Neutral | [2, 6] |
| B/P | 0.658 | 0.573 | [2, 6] |
| fu_p_ | 0.009 | 0.004 | [2, 6] |
| Absorption | |  |  |
| Absorption model | First order |  |  |
| fu_gut_ | 1 | 1 | [2, 6] |
| *f*_a_ | 0.82 |  | [2, 6] |
| *F*_g_ | 0.95 |  | [6] |
| *P*Caco-2(10^−6^ cm/s) | 4.70 |  | [6] |
| Reference Compound | Multiple |  | [6] |
| Scalar | 2.997 |  | [6] |
| Distribution | |  |  |
| Distribution model | Minimal PBPK model |  |  |
| *Q* (l/h) | 0.873 |  | [2, 6] |
| *V*_sac_ (l/kg) | 0.113 |  | [2, 6] |
| *V*_ss_ (l/kg) | 0.332 | 0.232 | [2, 6] |
| Elimination | |  |  |
| CL_iv_ (l/h) |  | 0.387 | [6] |
| CL_po_ (l/h) | 1.39 |  | [6] |
| rCYP3A4 CL_int_ (µL/min/pmol of isoform) | 0.175 | 0.141 | [2, 6] |
| Additional HLM clearance (µL/min/mg protein) | 7.359 | 4.819 | [2, 6] |
| Interaction | |  |  |
| CYP3A4 K_i_ (µM) | 12.5 | 11.65 | [6] |
| fu_mic_ | 0.876 | 0.730 | [6] |
| CYP2C9 K_i_ (µM) | 13.5 | 7.0 | [6] |
| fu_mic_ | 0.876 | 0.730 | [6] |
| CYP2C19 K_i_ (µM) | 22.5 | 19.55 | [6] |
| fu_mic_ | 0.876 | 0.730 | [6] |
| CYP2C8 K_i_ (µM) | 13 | 6.05 | [6] |
| fu_mic_ | 0.738 | 0.52 | [6] |
| CYP2CB6 K_i_ (µM) |  | 21.2 | [6] |
| fu_mic_ |  | 0.52 | [6] |
| CYP2D6 K_i_ (µM) |  | 24.4 | [6] |
| fu_mic_ |  | 0.730 | [6] |

B/P, blood-to-plasma ratio; CL_int_, intrinsic clearance; CL_iv_, in vivo intravenous clearance; CL_po_, in vivo oral clearance; *f*_a_, fraction absorbed; *F*_g_, fraction escaping gut-wall elimination; fu_gut_, fraction unbound in the enterocyte; fu_mic_, fraction unbound in the in vitro microsomal incubation; fu_p_, fraction unbound in plasma; HLM, human liver microsome; K_i_, concentration of inhibitor that supports half maximal inhibition; Log P_o:w_, logarithmic partition coefficient octonal:water; *Q*, inter-compartment clearance; *V*_sac_, single adjusted compartment volume; *V*_ss_, volume of distribution at steady state.

**Table S3.** Parameters Used to Develop the Elexacaftor Model in Simcyp**®** version 19 (Certara)

| **Parameter** | **Value** | **Source** |
| --- | --- | --- |
| Physiochemical properties | |  |
| Molecular weight (g/mol) | 597.66 | [2] |
| Log P_o:w_ | 6.00 | [2] |
| Compound type | Monoprotic acid | [2] |
| p*K*_a_ | 5.04 | [2] |
| B/P | 0.55 | [2] |
| fu_p_ | 0.00704 | [2] |
| Absorption | |  |
| Absorption model | First order |  |
| fu_gut_ | 7.75e-4 | [7] |
| *f*_a_ | 0.82 | [2] |
| *k*_a_ (h^−1^) | 0.59 | [2] |
| T_lag_ (h) | 2.17 | [2] |
| *P*Caco-2(10^−6^ cm/s) | 3.08 | [7] |
| Distribution | |  |
| Distribution model | Minimal PBPK model |  |
| *Q* (l/h) | 10.05 | [2] |
| *V*_sac_ (l/kg) | 0.27 | [2] |
| *V*_ss_ (l/kg) | 0.62 | [2] |
| Elimination | |  |
| CL_iv_ (l/h) | 1.25 | [7] |
| rCYP3A4 CL_int_ (µL/min/pmol of isoform) | 0.233 | [7] |
| Biliary CL_int_ (Hep) (µL/min/10^6^) | 4.56 | [2] |
| Interaction |  |  |
| CYP2C8 K_i_ (µM) | 8.35 | [7] |
| fu_mic_ | 0.399 | [7] |
| CYP2C9 K_i_ (µM) | 5.45 | [7] |
| fu_mic_ | 0.399 | [7] |

B/P, blood-to-plasma ratio; CL_int_, intrinsic clearance; CL_iv_, in vivo intravenous clearance; *f*_a_, fraction absorbed; fu_gut_, fraction unbound in the enterocyte; fu_mic_, fraction unbound in the in vitro microsomal incubation; fu_p_, fraction unbound in plasma; *k*_a_, absorption rate constant; K_i_, concentration of inhibitor that supports half maximal inhibition; Log P_o:w_, logarithmic partition coefficient octonal:water; p*K*_a_, logarithm of acid dissociation constant; *Q*, inter-compartment clearance; *T*_lag_, lag time; *V*_sac_, single adjusted compartment volume; *V*_ss_, volume of distribution at steady state.

*additional simulations were run modifying the fu_gut_ in published model to assess a value equal to 1 but it was unable to capture the clinically observed data so no modifications were made.

**Table S4.** Summary of the predicted Geometric Mean Ratio (90% CIs) for standard doses of elexacaftor (200mg q24h), tezacaftor (100mg q24h) and ivacaftor (150mg q12h) in the presence and absence of ritonavir (150mg q12h administered on day 1 through day 5)

| Drugs | | Predicted GMR (90% CI) of ETI PK parameters in the presence and absence of ritonavir on day 6 | |
| --- | --- | --- | --- |
| Substrate | Modulator | Cmax | AUC |
| Elexacaftor | Ritonavir | 2.02  (1.95, 2.10) | 2.31  (2.20, 2.42) |
| Tezacaftor |  | 2.18  (2.11, 2.25) | 3.11  (2.96, 3.27) |
| Ivacaftor |  | 6.84  (6.28, 7.45) | 9.31  (8.28, 10.47) |

**Table S5.** Predicted mean Cmax and AUC of reduced dose of ETI (one orange tablet q48h instead of two orange tablets in the morning and one blue tablet in the evening) with ritonavir (150 mg q12h) administered day 1 through day 5.

| Drug regimen with ritonavir administered day 1 through 5 | | | Cmax and  % of standard dose  ETI alone | | AUC and  % of standard dose  ETI alone | |
| --- | --- | --- | --- | --- | --- | --- |
| Drug Regimen | | Days | Cmax (mg/L) | % of ETI alone | AUC(0-48h) (mg∙h/L) | % of ETI alone |
| Elexacaftor  100mg q48h | With ritonavir | Day 1-3 | 6.9 | 85.2 | 282.8 | 89.5 |
|  |  | Day 3-5 | 6.6 | 81.5 | 272.8 | 86.3 |
|  | After ritonavir | Day 5-7 | 6.5 | 80.2 | 265.9 | 84.1 |
|  |  | Day 7-9 | 6.1 | 75.3 | 231.3 | 73.2 |
| Tezacaftor  50mg q48h | With ritonavir | Day 1-3 | 5.8 | 69.9 | 198.9 | 87.2 |
|  |  | Day 3-5 | 6.3 | 75.9 | 213.9 | 93.8 |
|  | After ritonavir | Day 5-7 | 6.5 | 78.3 | 219.7 | 96.3 |
|  |  | Day 7-9 | 6.3 | 75.9 | 180.9 | 79.4 |
| Ivacaftor  75mg q48h | With ritonavir | Day 1-3 | 1.2 | 75.0 | 49.9 | 93.0 |
|  |  | Day 3-5 | 1.7 | 106.3 | 68.8 | 128.3 |
|  | After ritonavir | Day 5-7 | 2.1 | 131.3 | 81.9 | 152.8 |
|  |  | Day 7-9 | 1.9 | 118.8 | 57.8 | 107.8 |

**Table S6.** Predicted mean Cmax and AUC of reduced dosing of ivacaftor (150mg 5 days apart) with ritonavir (150 mg q12h) administered day 1 through day 5.

| Ivacaftor 150mg 5 days apart | | Cmax and  % of standard dose  ivacaftor alone | | AUC and  % of standard dose  ivacaftor alone | |
| --- | --- | --- | --- | --- | --- |
| Ivacaftor dose | Days | Cmax (mg/L) | % of ivacaftor alone | AUC* (mg∙h/L) | % of ivacaftor alone |
| 150mg on day 1 | day 1-2 | 1.8 | 111.9 | 35.9 | 134.0 |
|  | day 1-6 |  |  | 151.5 | 113.0 |
| 150mg on day 6 | day 6-7 | 2.5 | 156.9 | 48.3 | 180.2 |
|  | day 6-9 |  |  | 101.4 | 126.1 |

*AUC(0-24h) for day 1-2 and day 6-7, AUC(0-120h) for day 1-6, and AUC(0-72h) for 6-9.

**Figure S1.** Sensitivity analysis of the fmCYP3A4 of ivacaftor on the predicted AUC ratio with or without ketoconazole (A) or rifampin (B). The grey-colored area shows the 90% confidence interval of predicted AUC ratio.

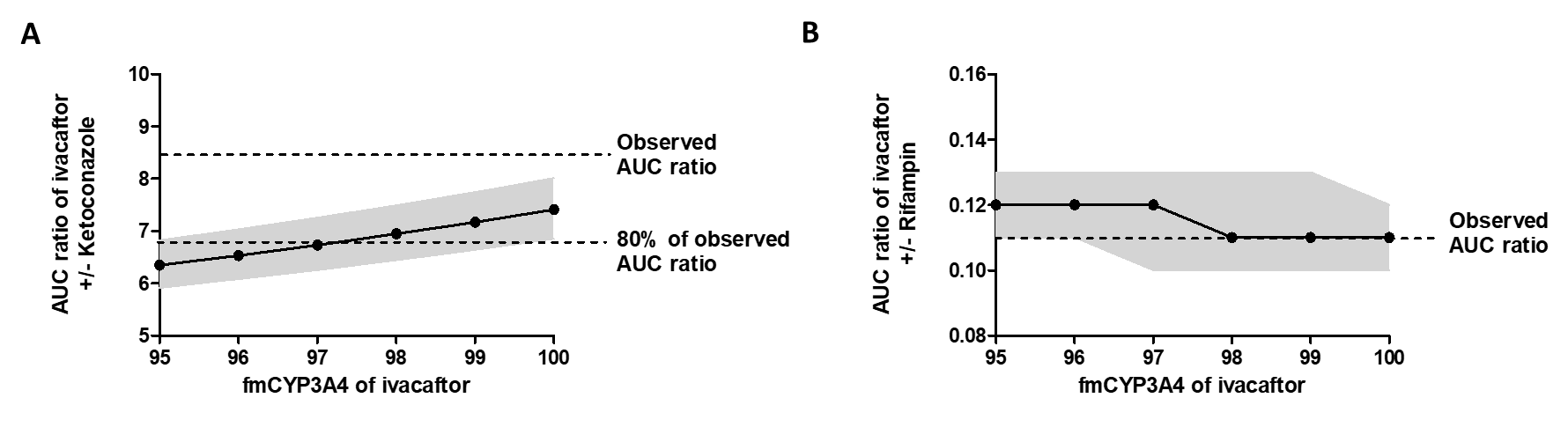

**Figure S2.** Observed and simulated plasma concentration-time profiles of ETI following a single oral dose of elexacaftor 200mg (A), tezacaftor 100mg (B), and ivacaftor 100mg (C). The grey-colored area shows the range of predicted concentrations from 95^th^ to 5^th^ percentiles.

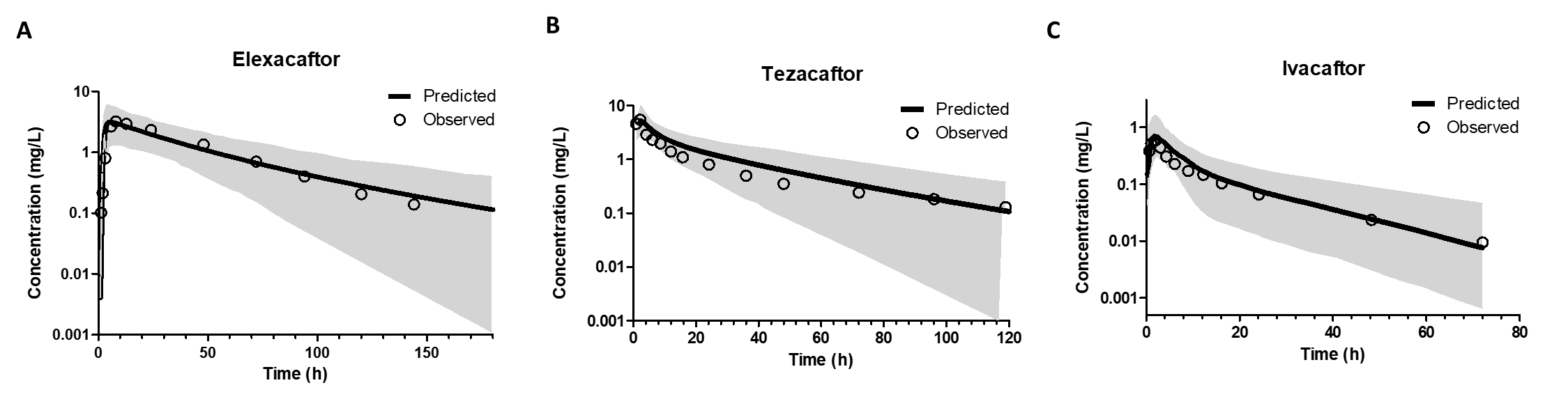

**Figure S3**. Plasma Concentration Profile of ETI. Green: standard dose without ritonavir, Red: reduced dose (elexacaftor 100mg q48h, tezacaftor 50mg q48h, ivacaftor 75mg q48h) with ritonavir 150mg q12h administered day 1 through day 5

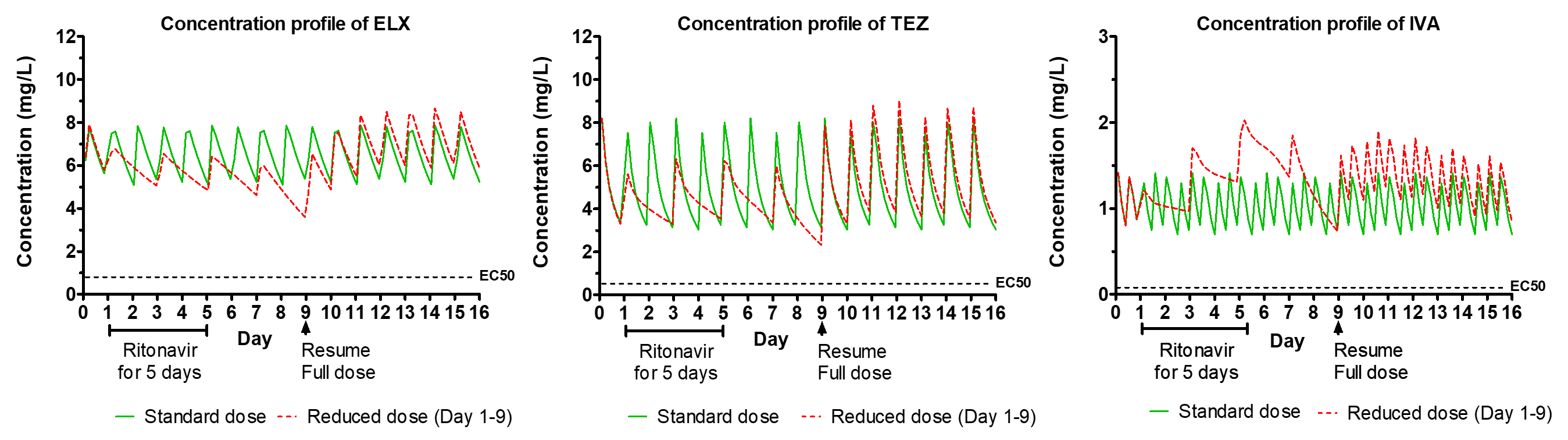
